# Leveraging transformers and large language models with antimicrobial prescribing data to predict sources of infection for electronic health record studies

**DOI:** 10.1101/2024.04.17.24305966

**Authors:** Kevin Yuan, Chang Ho Yoon, Qingze Gu, Henry Munby, A Sarah Walker, Tingting Zhu, David W Eyre

**Affiliations:** Big Data Institute, Nuffield Department of Population Health, University of Oxford, Oxford, UK; Nuffield Department of Medicine, University of Oxford, Oxford, UK; University Hospitals Bristol & Weston NHS Trust, Bristol, UK; NIHR Health Protection Research Unit in Healthcare Associated Infections and Antimicrobial Resistance, University of Oxford, Oxford, UK; NIHR Oxford Biomedical Research Centre, Oxford, UK; Institute of Biomedical Engineering, University of Oxford, Oxford, UK; Oxford University Hospitals NHS Foundation Trust, Oxford, UK

## Abstract

**Background:** Electronic health records frequently contain extensive unstructured free-text data, but extracting information accurately from these data and at scale is challenging. Using free-text from antibiotic prescribing data as an example, we investigate the performance of modern natural language processing methods (NLP) and large language models (LLMs) as tools for extracting features from medical records.

**Methods:** We used 938,150 hospital antibiotic prescriptions from Oxfordshire, UK. The 4000 most frequently used free-text indications justifying antibiotic use were labelled by clinical researchers into 11 categories describing the infection source/clinical syndrome being treated and used for model training. Traditional classification methods, fuzzy regex matching and n-grams with XGBoost, were compared against modern transformer models: we fine-tuned generic and domain-specific BERT models, fine-tuned GPT3.5, and investigated few-shot learning with GPT4. Models were evaluated on internal and external test datasets (2000 prescriptions each). Infection sources determined from ICD10 codes were also used for comparisons.

**Results:** In internal and external test datasets, the fine-tuned domain-specific Bio+Clinical BERT model averaged an F1 score of 0.97 and 0.98 respectively across the classes and outperformed the traditional regex (F1=0.71 and 0.74) and n-grams/XGBoost (F1=0.86 and 0.84). OpenAI’s GPT4 model achieved F1 scores of 0.71 and 0.86 without using labelled training data and a fine-tuned GPT3.5 model F1 scores of 0.95 and 0.97. Comparing infection sources extracted from ICD10 codes to those parsed from free-text indications, free-text indications revealed 31% more specific infection sources.

**Conclusion:** Modern transformer-based models can efficiently and accurately categorise semi-structured free-text in medical records, such as prescription free-text. Finetuned local transformer models outperform LLMs currently for structured tasks. Few shot LLMs match the performance of traditional NLP without the need for labelling. Transformer-based models have the potential to be used widely throughout medicine to analyse medical records more accurately, facilitating better research and patient care.

## Introduction

Electronic health records (EHRs) offer unprecedented quantities of structured and unstructured data for driving research and improving care delivery. Manually extracting relevant information from unstructured free-text EHRs is costly and laborious. Recent developments in natural language processing (NLP) and the advent of large language models (LLMs) offer promising and potentially transformational alternatives that can accurately acquire relevant information from unstructured text for patient diagnosis, ^1–3^ as well as perform several routine tasks from medical records.^4,5^

As in other medical domains, studies of antibiotic resistance, use, and stewardship have traditionally relied on manual review of clinical notes and prescriptions^6–8^ or mapping of International Classification of Diseases (ICD) diagnostic codes to identify acute infection diagnoses and chronic comorbidities.^9,10^ However, in studies of sepsis, ICD codes were less sensitive than clinical data for detecting cases,^11,12^ particularly for less common infections like meningitis,^13^ and had variable validity.^14^ Additionally, since codes are recorded only after patient discharge, assigned infection sources may not align with individual antibiotic prescriptions, particularly during the more uncertain initial phase of inpatient care. Conversely, manual chart review has higher sensitivity and can detect indications evolving over time, but time and cost constraints mean that case numbers are often limited.

For research, applying LLMs to entire medical records can effectively make predictions and generate new medical content.^4,5^ However, there is also a clear need for research and service applications to be able to extract specific individual features from EHRs reliably and efficiently whilst also meeting information governance requirements. As an example of this targeted approach, we investigated several methods using electronic prescribing data to classify the source of an infection/infectious syndrome being treated, analysing free-text indications documented by clinicians^15^ justifying the reason for antibiotic prescriptions. We compared state-of-the-art NLP models, Bidirectional Encoder Representations from Transformers (BERT),^16^ LLMs, and Generative Pretrained Transformer (GPT),^17^ to traditional ICD code-based approaches, classical NLP methods and regular expression-based text searches. We show modern approaches have potential to accurately extract key information from medical records at scale, potentially opening opportunities for new epidemiological and intervention studies across all of medicine, as well as possibilities for improving care delivery.

## Methods

### Study design and population

We used EHRs from two distinct hospital sites, Oxford and Banbury, with Oxford serving as our training and internal test set, and Banbury as our external test set. These sites collectively provide 1100 beds, serving 750,000 residents in Oxfordshire, ∼1% of the UK population. Deidentified data were obtained from Infections in Oxfordshire Research Database (IORD), which has approvals from the National Research Ethics Service South Central – Oxford C Research Ethics Committee (19/SC/0403), the Health Research Authority and the Confidentiality Advisory Group (19/CAG/0144) as a deidentified database without individual consent. All patients aged ≥16 years who had antibiotic prescriptions were included.

### ‘Ground Truth’ Labelling

Two clinical researchers reviewed each antibiotic indication text string used for training and testing (described below) to establish a reference or ‘ground truth’ label for the clinical syndrome being treated. Any discrepancies were resolved by a third researcher, a clinical infection specialist. Antibiotic indications were labelled using 11 categories representing infection source: Urinary; Respiratory; Abdominal; Neurological; Skin and Soft Tissue; Ear, Nose and Throat (ENT); Orthopaedic; Other specific (i.e. another body site); Non-specific (i.e. no body site provided, e.g. “sepsis”), Prophylaxis, Not informative (i.e. text unrelated to the source of infection, e.g. “as instructed by Dr X”). Each category was recorded as a binary variable, such that more than one potential source could be recorded, e.g. the input string “urinary/chest” would be labelled as both urinary and respiratory. An additional variable was used to document the presence of uncertainty expressed by the prescriber, e.g. “urinary/chest” or “? UTI”.

### Comparator Classification by ICD10 Codes

As a comparator, we mapped primary and secondary ICD10 diagnosis codes from the same admission as the antibiotic prescription to the 11 infection sources using CCSR classifications^18^ as an intermediate step (see Supplement and *Appendix S1*).

### Traditional Classification Methods

#### Regex Rules

The most intuitive and deterministic method for classifying free-text is searching for specific keywords from a list of predefined words for a given category. We employed fuzzy regular expression (regex) matching patterns with term-specific word boundaries and variable fuzziness to allow for misspellings and variations (see Supplement and *Appendix S2*).

#### n-grams & XGBoost

A second approach used a separate tokeniser, embedding and classifier structure; specifically, Scikit-learn’s n-grams & count vectorisation and the gradient boosting model architecture XGBoost.^19,20^ Each free-text indication term was broken up into overlapping subwords of length $n$ and then count-vectorised, with the count representing the frequency of each subword’s occurrence. The vectors of dimension $vocabulary size$ were then fed as input features to the classification model. We determined the optimal n-gram size ($n$) and hyperparameters for XGBoost during model training by maximising the receiver operator curve area under the curve (ROC-AUC) (details below).

### BERT Classifier

Current state-of-the-art NLP tasks employ models built on Transformer architectures, with the Bidirectional Encoder Representations from Transformers (BERT) model family well suited for many tasks requiring semantic understanding. We finetuned^21^ pre-trained BERT models on a single GPU instance and used BERT for both encoding and classification. We evaluated the original generic “uncased base BERT” model, pre-trained on the BooksCorpus and English Wikipedia and a domain-specific “Bio+Clinical BERT”, pre-trained on biomedical and clinical text.^22,23^

### Few-Shot and finetuned LLM Classifier

Compared to BERT, the GPT family enables zero- or few-shot learning, i.e. there is potentially minimal need for labelled data for task-specific training. We developed prompts for GPT4, comprised of instructions and the target categories, asking the model to complete the categories (see *Appendix S3* for specific prompts, and *Appendix S4* for details of batch sizes).^24^

Similarly, a GPT3.5 model was finetuned on the training dataset and instructed to classify the free-text. Finetuning was achieved by presenting the desired output alongside the training input data. We used the same system prompt as for the few-shot model, while providing the training examples which were fed in batches of ten. Additional model hyperparameters, such as learning rate, and epochs, were chosen through grid search.

### Training, Test and External Evaluation

We divided the Oxford data with a 90/10 train/test split, resulting in a raw training set and internal test set. From the training data, we labelled and used the 4000 most frequently occurring unique indications. To make labelling tractable we discarded the remaining unlabelled data from the training set. From the internal test data, we randomly selected and exhaustively labelled indications present in 2000 prescriptions. For the external test set from Banbury, we also labelled 2000 randomly selected entries. All models were trained on the training dataset with grid-search based hyperparameter tuning based on cross validation and tested on both the internal and external test sets.

The multi-label performance of each method was evaluated using weighted F1 scores, precision-recall (PR-AUC) and ROC-AUC. Weighted averages take into account the varied distributions of the classes, such that more common classes contribute more to the overall average, producing estimates that reflect the original data source and represent real-world performance.

## Results

We obtained antibiotic prescribing indication data from 826,533 prescriptions from 171,460 adult inpatients, ≥16 years, between 01-October-2014 and 30-June-2021 from three hospitals in Oxford, UK. The most commonly prescribed antibiotics were co-amoxiclav (n=269,945, 33%), gentamicin (n=70,002, 8%), and metronidazole (n=65,094, 8%) (*Appendix S5*), and the most common specialities were General Surgery (n=146,719, 18%), Acute General Medicine (n=98,687, 12%), and Trauma and Orthopaedics (n=90,719, 11%) (*Appendix S6*). Patients were a median 56 years old (IQR 36-73), and 94,721 (55%) were female.

We also used an independent external test dataset to assess classifier performance further, from the Horton Hospital, Banbury (∼30 miles from Oxford). This dataset comprised 111,617 prescriptions from 25,924 patients between 01-December-2014 and 30-June-2021, with 13,650 unique free-text indications. Antibiotics prescribed (*Appendix S5*) and specialities (*Appendix S6*) were broadly similar to the Oxford training set. Patients were a median 67 years old (IQR 47-80), and 13,853 (53%) were female.

### Prescription indications

From the 826,533 Oxford prescriptions, 86,611 unique free-text indications were recorded. The top 10 accounted for 41% of all prescriptions; these included “Perioperative Prophylaxis” (20%), “UTI” (4%), “LRTI” (3%), “Sepsis” (3%), and “CAP” (3%). The most commonly occurring 4000 unique indications, used for model training, accounted for 84% (692,310) of prescriptions (*Appendix S7*).

As expected, different wording was used to reflect similar concepts, e.g. “CAP [community acquired pneumonia]”, “LRTI [lower respiratory tract infection]”, “chest infection”, and “pneumonia”. Additionally, misspellings were common, e.g. “infction”, “c. dififcile”. Multiple examples expressed uncertainty, or multiple potential sources of infection, e.g. “sepsis ?source”, “UTI/Chest”, etc. Reflecting the complexity of prescribing, there were multiple potentially informative, but rarely occurring indications, e.g., “transplant pyelonephritis”, “Ludwig’s angina”, and “deep neck infection”, which were only seen 51 (<1%), 27 (<1%), and 13 (<1%) times respectively (*Appendix S8*).

### ‘Ground truth’ labels

Following labelling by clinical experts, the 4000 most commonly occurring free-text indications were classified into 11 categories, with a separate variable capturing the presence of uncertainty. The most commonly assigned sources were “Prophylaxis” (267,788/692,310 prescriptions, 39%), “Respiratory” (125,744, 18%) and “Abdominal” (61,670, 9%). 50% (n=344,773) prescriptions had “No Specific Source”. The most uncertainty was expressed in “Neurological” and ENT cases at 38% and 33%, respectively (*Figure 1A*). Although “Respiratory” was the most common category overall after “Prophylaxis”, there were more distinct text strings associated with “Abdominal” infections, with “Skin and Soft Tissue” infection also having a disproportionately larger number of unique text strings (*Figure 1B*). Most ‘multi-source’ prescriptions were a combination of “Prophylaxis” and a source (>90%). Excluding prophylaxis, the most common combinations of sources were “No Specific source” and “Not Informative”, “Urinary” and “Respiratory”, and “Skin & Soft Tissue” and ENT, in 1.6%, 0.58%, 0.41% prescriptions, respectively (*Figure 1C-D*). The former two reflected diagnostic uncertainty and the latter reflected infections of the face, head and neck frequently involving skin/soft tissue.

**Figure 1:**
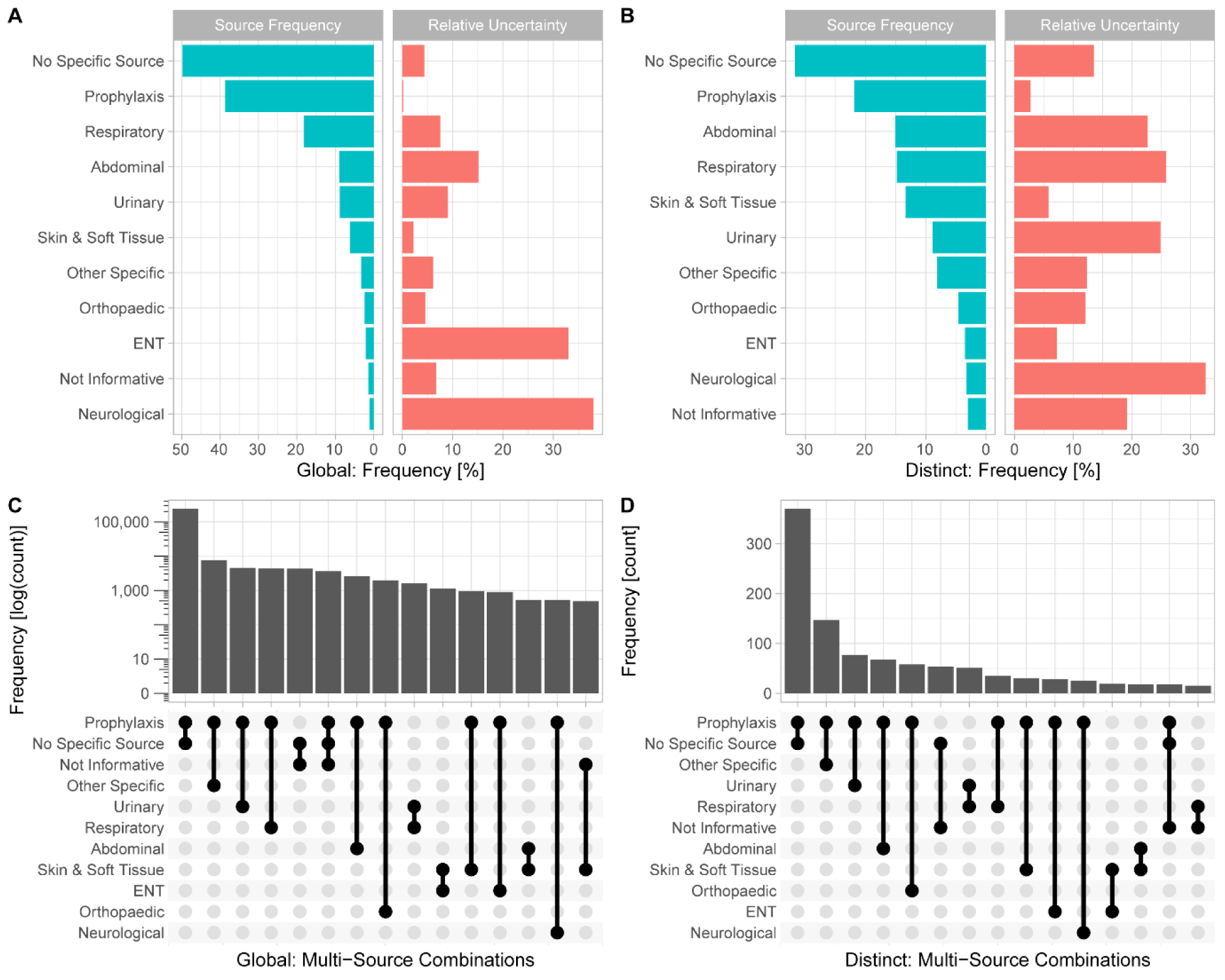
Infection source distributions within labelled training data from three Oxford hospitals. *Bar charts A) and B) show the distribution of the sources and the uncertainty relative to the infection source. The up-set plots C) and D) show the occurrence of multiple sources within the same prescription. Panels A) & C) show distributions across the entire labelled indications training set, panels B) & D) across a distinct set of 4000 most common indications. *Indications falling into ENT such as “neck abscess” were often also labelled with Skin & Soft Tissue.*

### Classifier performance

We trained classifiers using the labelled training data from three Oxford hospitals (*Figure 2*). Compared to clinician-assigned labels, within the internal Oxford test dataset (n=2000), the weight-averaged F1 score across classes was highest using Bio+Clinical BERT (Average F1=0.97 [worst performing class F1=0.84, best performing F1=0.98]) followed by finetuned GPT3.5 (F1=0.95 [0.77-0.99]), base-BERT (F1=0.93 [0.23-0.98]) and tokenisation+XGBoost (F1=0.86 [0.64-0.96]). Nearly all approaches exceeded traditional regular expression-based matching (F1=0.71 [0.00-0.93]). The few-shot GPT4 model which did not require labelled data performed similarly to this baseline (F1= 0.71 [0.30-0.98]). Similar performance characteristics were achieved on the external validation dataset from Banbury (n=2000; weight-averaged F1 scores: Bio+Clinical BERT 0.98 [0.87-1.00], finetuned GPT3.5 0.97 [0.70-1.00], Base BERT 0.97 [0.63-0.99], XGBoost 0.84 [0.63-1.00], Regex 0.74 [0.00-0.96], GTP4 0.86 [0.25-1.00]) (Table 1, also shows classification run times).

**Figure 2:**
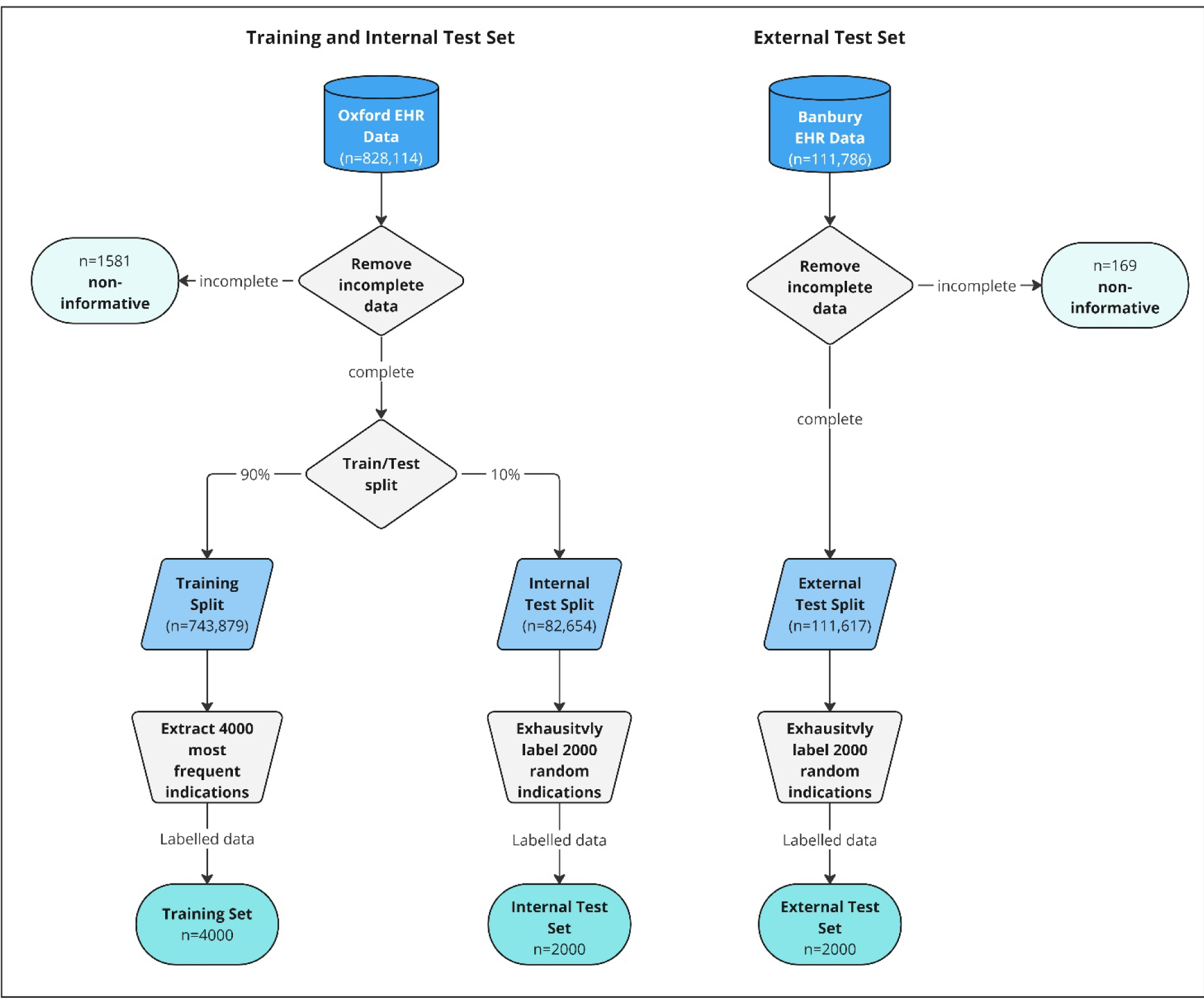
Data processing flow chart for training and internal and external test data sets. *Prescribing data was fetched from EHR databases and filtered for complete data. The 4000 most frequent indications within the training split were labelled, all remaining training data was discarded. 2000 entries were randomly sampled from both the internal and external test datasets and exhaustively labelled, resulting in a total of three datasets (training set, internal test set, external test set)*.

**Table 1:**
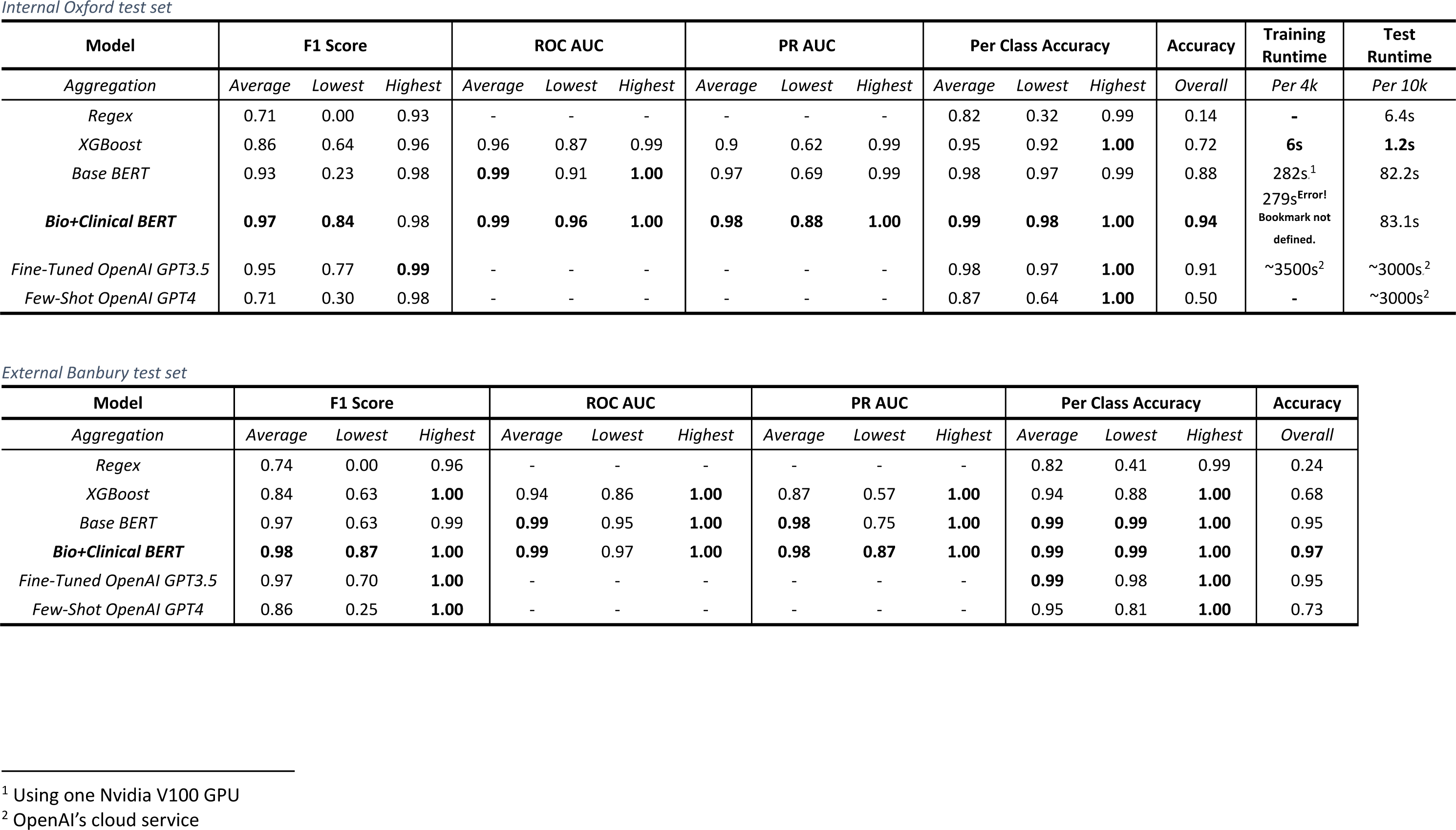
Model performance metrics for the internal (Oxford) and external (Banbury) test sets. *Each score is listed with the weighted average across the classes (sources), with the lowest and highest performing class. Overall Accuracy refers to the score calculated for a sample treated as a whole. The finetuned Bio+Clinical BERT outperforms all other methods on both internal and external test sets*.

| Model | F1 Score |  |  | ROC AUC |  |  | PR AUC |  |  | Per Class Accuracy |  |  | Accuracy | Training Runtime | Test Runtime |
| --- | --- | --- | --- | --- | --- | --- | --- | --- | --- | --- | --- | --- | --- | --- | --- |
| Aggregation | Average | Lowest | Highest | Average | Lowest | Highest | Average | Lowest | Highest | Average | Lowest | Highest | Overall | Per 4k | Per 10k |
| Regex | 0.71 | 0.00 | 0.93 | - | - | - | - | - | - | 0.82 | 0.32 | 0.99 | 0.14 | - | 6.4s |
| XGBoost | 0.86 | 0.64 | 0.96 | 0.96 | 0.87 | 0.99 | 0.9 | 0.62 | 0.99 | 0.95 | 0.92 | **1.00** | 0.72 | **6s** | **1.2s** |
| Base BERT | 0.93 | 0.23 | 0.98 | **0.99** | 0.91 | **1.00** | 0.97 | 0.69 | 0.99 | 0.98 | 0.97 | 0.99 | 0.88 | 282s<sup>1</sup> | 82.2s |
| **Bio+Clinical BERT** | **0.97** | **0.84** | 0.98 | **0.99** | **0.96** | **1.00** | **0.98** | **0.88** | **1.00** | **0.99** | **0.98** | **1.00** | **0.94** | 279s<sup>Error!</sup><br>Bookmark not defined. | 83.1s |
| Fine-Tuned OpenAI GPT3.5 | 0.95 | 0.77 | **0.99** | - | - | - | - | - | - | 0.98 | 0.97 | **1.00** | 0.91 | ~3500s<sup>2</sup> | ~3000s<sup>2</sup> |
| Few-Shot OpenAI GPT4 | 0.71 | 0.30 | 0.98 | - | - | - | - | - | - | 0.87 | 0.64 | **1.00** | 0.50 | - | ~3000s<sup>2</sup> |

External Banbury test set
| Model | F1 Score |  |  | ROC AUC |  |  | PR AUC |  |  | Per Class Accuracy |  |  | Accuracy |
| --- | --- | --- | --- | --- | --- | --- | --- | --- | --- | --- | --- | --- | --- |
| Aggregation | Average | Lowest | Highest | Average | Lowest | Highest | Average | Lowest | Highest | Average | Lowest | Highest | Overall |
| Regex | 0.74 | 0.00 | 0.96 | - | - | - | - | - | - | 0.82 | 0.41 | 0.99 | 0.24 |
| XGBoost | 0.84 | 0.63 | **1.00** | 0.94 | 0.86 | **1.00** | 0.87 | 0.57 | **1.00** | 0.94 | 0.88 | **1.00** | 0.68 |
| Base BERT | 0.97 | 0.63 | 0.99 | **0.99** | 0.95 | **1.00** | **0.98** | 0.75 | **1.00** | **0.99** | **0.99** | **1.00** | 0.95 |
| **Bio+Clinical BERT** | **0.98** | **0.87** | **1.00** | **0.99** | 0.97 | **1.00** | **0.98** | **0.87** | **1.00** | **0.99** | **0.99** | **1.00** | **0.97** |
| Fine-Tuned OpenAI GPT3.5 | 0.97 | 0.70 | **1.00** | - | - | - | - | - | - | **0.99** | 0.98 | **1.00** | 0.95 |
| Few-Shot OpenAI GPT4 | 0.86 | 0.25 | **1.00** | - | - | - | - | - | - | 0.95 | 0.81 | **1.00** | 0.73 |

<sup>1</sup> Using one Nvidia V100 GPU
<sup>2</sup> OpenAI’s cloud service

### Classifier Performance by Class

Using the best performing classifier, Bio+Clinical BERT, we assessed performance within each category. The best-performing categories within our internal test set were “Respiratory”, “No Specific Source” and “Prophylaxis” (F1 score=0.98), followed by “Urinary” (0.97), “Abdominal” (0.96), “Orthopaedic” (0.90), “Not Informative” (0.89) and “Neurological” (0.88). The worst performing category was Orthopaedic (0.84), likely due to the high variety of terms used and low number of training samples (n=14, *Appendix S9*). Uncertainty was also well detected (0.96) (*Figure 3A, Appendix S10*).

**Figure 3:**
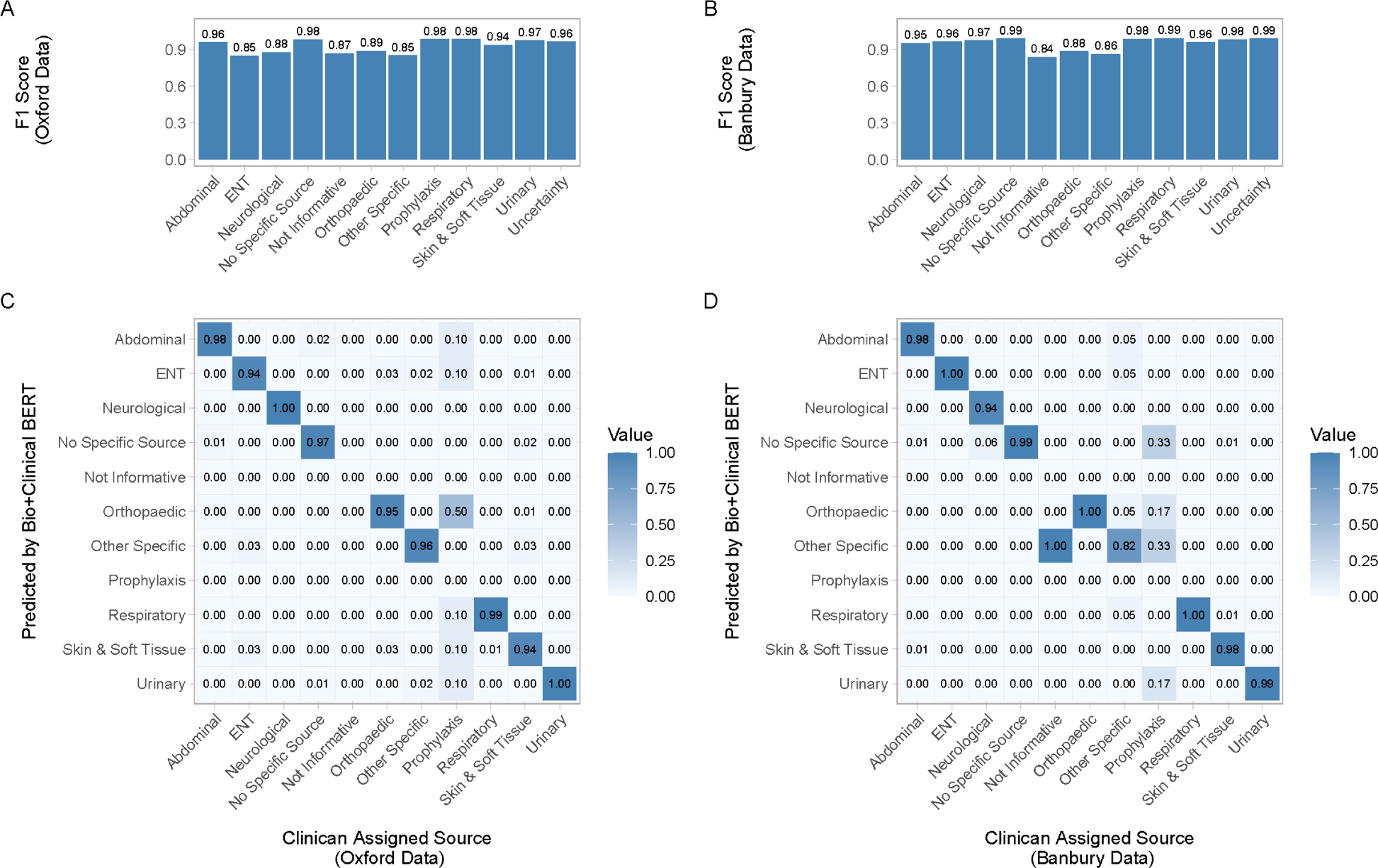
Performance metrics for Bio+Clinical BERT on both internal and external test sets. *Bar charts A) and B) show the per-class prediction performance. Confusion matrices C) and D) are single indication test cases and show model prediction errors across the sources for given ground truths (clinician assigned sources). Panels A) and C) show evaluations performed on the internal test set from three Oxford hospitals, B) and D) on the external test set from the Banbury hospital*.

In the external test data, scores varied slightly, with all source categories except for “Not Informative” having F1 scores on average 0.02 higher compared to the internal test set. These small differences likely arose from different compositions of categories and the amount of shared vocabulary between the training and test datasets.

### Misclassifications

Most misclassifications were spread evenly across classes for single indications. The two most common misclassifications occurred for “Orthopaedic” and “Other Specific” cases, with 12% being misclassified as “Prophylaxis” and 8% as “Skin and Soft Tissue”, respectively on the internal test set. On the external test set, most misclassifications were predicted to be “Other Specific” or “Prophylaxis” (*Figure 3C*).

### Training Dataset Size

We examined the effect of training size on model performance using randomly selected training dataset subsets of 250, 500, 750, 1000, 1500, 2000, 3000, and 4000 unique indications, tested using both internal/external test sets. There was a notable increase in performance (AUC-ROC and F1 scores) when the training size increased from 250 to 1000 samples, suggesting a minimum of 1000 training samples for adequate performance. However, we saw limited improvement as training dataset size rose to 4000, indicating there may be only marginal gains to expanding the training data beyond 4000 samples (*Appendix S11*).

### Comparing free-text indications to ICD10 codes

We also compared infection sources from manually labelled ‘ground truth’ free-text indications to sources inferred from ICD10 diagnostic codes. 31% of sources classified as “unspecific” using diagnostic codes could be resolved into specific sources using free-text. Rarer infection sources such as “CNS” and “ENT” (<1% and no occurrence in diagnostic codes) were represented better by sources extracted from free-text (4% and 4% respectively). Overall, where defined, sources listed in clinical codes generally concurred with the ‘ground truth’ free-text sources (*Figure 4*).

**Figure 4:**
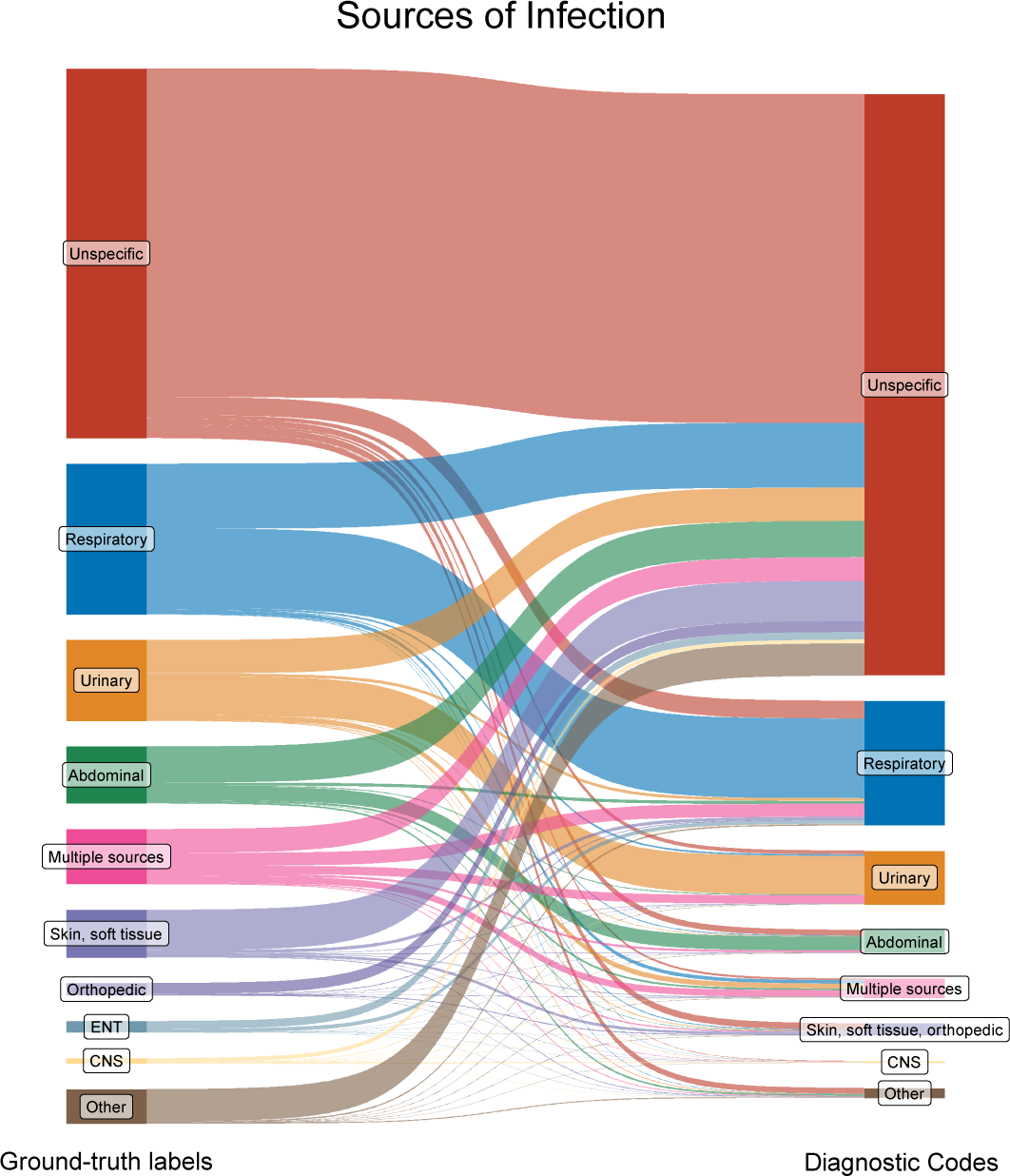
Comparing infection sources between clinician assigned free-text indications (left) and diagnostic codes (right) in the training and internal and external test sets. *Clinician assigned categories were extracted from prescribing data and manually labelled, diagnostic codes sources calculated from procedure and discharge codes*.

## Discussion

We show that modern NLP methods can extract clinically relevant details from semi-structured free-text fields. In our example application, a finetuned Bio+Clincal BERT transformer model classified the infection source being treated using clinician-written antibiotic indication text with an F1 score of 0.97 (harmonic mean of sensitivity and positive predictive value). Although this required manual labelling of several thousand text strings, exhaustive labelling of all possible prior strings was not required to achieve this performance. The accuracy of the Bio+Clinical BERT model was substantially better than a sophisticated regular expression-based approach, despite the latter being the solution that many healthcare institutions and researchers might choose at present.

We also explored what performance might be possible using LLMs without having to label data, e.g. where this is not possible or too resource intensive. However, few-shot learning with GPT4 only achieved modest performance, but it was still similar to the baseline regex method and more consistent across classes. Using LLMs with labelled training data, i.e. a fine-tuned GPT3.5 variant achieved results comparable to the Bio+Clinical BERT approach when correctly specified and tuned but showed more limited potential for deployment as responses can vary greatly in formatting, making it difficult to parse correctly into a rigid format required for most downstream tasks or EHR systems. In environments with limited computing resources where the deployment of deep-learning models is not feasible, regex and XGBoost-based models provide possible alternatives with a reduced runtime of 6.4 and 1.2 seconds/10k indications vs 82.2 seconds/10k for Bio+Clinial BERT.

Currently, research or clinical use of free-text may be limited by concerns that personal data may be included. Here, by homogenising and categorising sensitive free-text data, we present a privacy-aware solution that enables researchers to utilise the depth of free-text data without direct access or the possibility of identifying specific patients.

Our study has several limitations, including that we only used a subset of the available training data, through the non-exhaustive labelling of a subset of antibiotic indication text strings. However, labelling the 4000 most common unique terms, accounting for 84% of the data, achieved very high performance, with sensitivity analyses suggesting that labelling more examples would not have improved performance substantially. This is likely possible because the underlying Bio+Clinical BERT model is already pre-trained on medical terms and capable of inferring similar words. Of note, many of the remaining 17% of text strings were different combinations of already labelled words, suggesting fewer “new” or unseen keywords than might be expected. Not fully labelling the training data also makes it more difficult to compare class distributions with the test data sets. We also only used a subset of the test data to evaluate performance; however the 2000 randomly selected samples are likely representative. Although the labelling process was somewhat subjective, independent labelling by two clinical researchers with a third adjudicating any discrepancies was designed to minimise this.

Future enhancements could optimise computational efficiency while maintaining comparable performance. While XGBoost offers faster training and inference times, its performance was poorer than Bio+Clinical BERT. Therefore, smaller, more efficient NLP models might balance computational demands and performance. Techniques such as model pruning, quantisation, and knowledge distillation could reduce model size and computational requirements while preserving performance.^25–27^ While GPT4 deployments can comply with data governance requirements, its use presents challenges in some settings, as it is usually accessed via third-party cloud compute providers rather than healthcare institutions. Where data need to remain on site, open-source, locally-deployed language models, such as LLAMA, ALPACA or Mistral 7B, may be alternatives that could be further investigated.^28–30^

Our approach has several possible applications; for example, it could be used to monitor and evaluate prescribing practice across different conditions, it provides classification of possible infection sources for epidemiological research,^31^ and is also a mechanism for extracting standardised features from medical records for use in predictive algorithms being developed to improve patient care. Although we demonstrate excellent performance for antibiotic indications, it could also be applied to other short strings of free-text, for example descriptions of surgical procedures, patient functional states, or presenting complaints in emergency department and hospital admission data. Across all these domains, refined patient stratification could improve both research and care delivery.

In summary, we show that state-of-the-art NLP can be used to efficiently and accurately categorise semi-structured free-text in medical records. This has the potential to be applied widely to analyse medical records more accurately.

## Data Availability

The data analysed are available from the Infections in Oxfordshire Research Database (https://oxfordbrc.nihr.ac.uk/research-themes/modernising-medical-microbiology-and-big-infection-diagnostics/infections-in-oxfordshire-research-database-iord/), subject to an application and research proposal meeting on the ethical and governance requirements of the Database. Labelled training and test datasets and the pre-trained BERT model are also available via an application to the Database.

https://github.com/kevihiiin/EHR-Indication-Processing/

## Acknowledgements and funding

This work was supported by the National Institute for Health Research Health Protection Research Unit (NIHR HPRU) in Healthcare Associated Infections and Antimicrobial Resistance at Oxford University in partnership with the UK Health Security Agency (NIHR200915), and the NIHR Biomedical Research Centre, Oxford. DWE is a Big Data Institute Robertson Fellow. ASW is an NIHR Senior Investigator. The views expressed are those of the authors and not necessarily those of the NHS, the NIHR, the Department of Health or the UK Health Security Agency. KY is supported by the EPSRC Centre for Doctoral Training in Health Data Science (EP/S02428X/1). The funders had no role in study design, data collection and analysis, decision to publish, or preparation of the manuscript.

## Declaration of interests

No other author has a conflict of interest to declare.

## Code availability

All tools developed for this study (Regex builder, Bio+Clinical BERT pipeline, GPT3.5 finetuning and GPT4 few-shot request tools), pre-trained models, as well as the evaluation framework, are available on GitHub:

https://github.com/kevihiiin/EHR-Indication-Processing/

## Supplementary appendix

### Supplementary methods

#### Comparator Classification by ICD10 Codes

We first summarised all ICD10 codes into broader concepts using the CCSR classification tool, grouping >70,000 ICD10 codes into 544 categories. Two independent clinical researchers then developed a custom lookup table mapping CCSR categories to our 11 infection source groups (*Appendix S1*), with a third clinician resolving discrepancies.

#### Traditional Classification Methods

##### Regex Rules

The regex patterns for each category were built using the 50 most common indications for that infection source in the training data, with individually assigned error-rate thresholds for inexact matching and specified word boundaries for each string. This allowed for strict exact matching on abbreviations while permitting spelling mistakes for longer words. Word boundaries ensured that abbreviations were not matched when part of a longer word (e. g. avoiding finding UTI in roUTIne post-op, cUTIbacterium). Our tool, designed to automate the creation of complex regex queries based on a given reference set and user specifications, can be found in *Appendix S*2. The assisted regex builder simplified creating complex matching rules for infection sources, extracting the most common indications for each category and exporting them into a pre-populated table with parsing options for additional user input. Users can then modify these rules: adding word boundaries for precise matches, setting error rates (e.g., zero for abbreviations), and excluding redundant words. The edited table is then read back and converted into complex regex-matching strings for each category. This allows for medical experts to build and modify complex matching rules without needing to understand and debug error-prone regular expressions.

#### BERT Classifier

We evaluated the performance of the original generic “uncased base BERT” model, pre-trained on the BooksCorpus and English Wikipedia and a domain-specific “Bio+Clinical BERT”, pre-trained on biomedical and clinical text sourced from PubMed, PubMed Central and MIMIC-III v1.4 notes.^22,23^ Both pre-trained models were fetched from the HuggingFace model hub (uploaded on the 18-June-2019 and 28-February-2020), and finetuned using the HuggingFace “transformers” library with indications as input and the source categories as output.^21^

#### Few-Shot and finetuned LLM Classifier

We developed prompts for GPT4, comprised of instructions and the target categories, asking the model to complete the categories (specific prompt in *Appendix S3*).^24^ We made several iterations to the prompt on a subset of the training data, aiming to increase the model’s understanding of the task. Given the model’s generative nature, we accessed GPT4 through the API and supplied inputs to create more structured, deterministic and less creative answers. Specifying a rigid output format is crucial for a multi-label task. We therefore instructed the model to present its prediction output in JSON format, using the original indication as the key and the categories as a list of values. To prevent the model from creating new categories, we penalised it for returning new tokens not seen in the text (i.e. the prompt) by setting a higher ‘presence penalty’. A fixed ‘seed’ and lower ‘temperature’ were chosen to coerce the model into returning more deterministic and reproducible answers.^32^ The same hyperparameters were used for prediction with the fine-tuned GPT3.5 model, aiming to increase determinism and reproducibility.

### Appendix S1 Code and LUTs Repository

**ICD10 to Infection Source Mapping Table**

*https://github.com/kevihiiin/EHR-Indication-Processing/blob/main/00_Data/LUTs/icd10_ccsr_mapping.csv*

### Appendix S2 Regex Rule Builder

**Figure S2:**
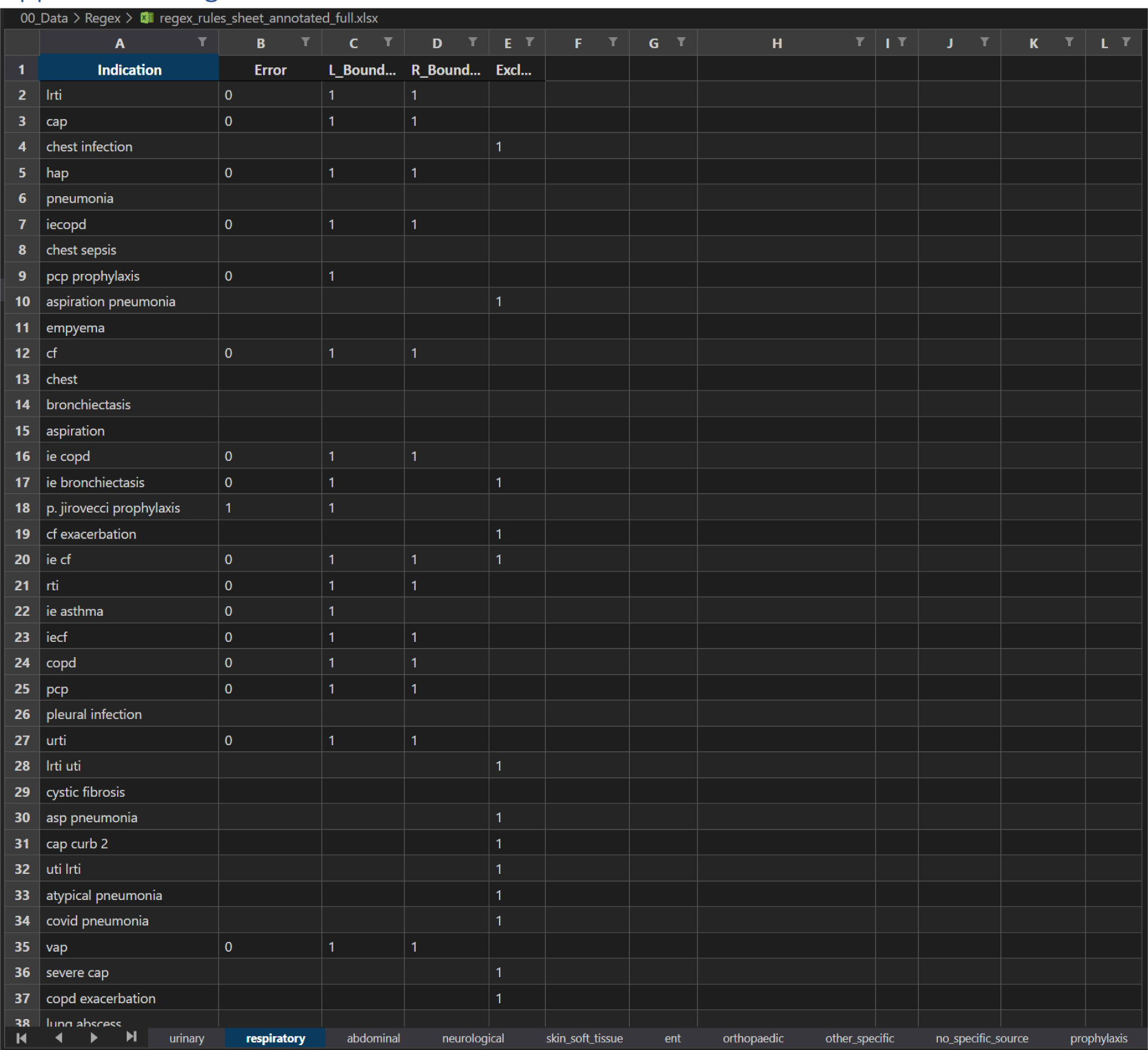
Regex Rule Builder user specifications. *Specify a non-default error rate for abbreviations, set left and right word boundaries and exclude duplicated words. One sheet per category*.

### Appendix S3 GPT4 Prediction Prompt (API)

*System prompt:*

*You are a helpful and precise UK medical expert; you have been given a list of indications describing why antibiotics were prescribed to patients in a hospital. You have been asked to **label** these indications into categories.*

*You can only **choose** from these categories which are: Urinary, Respiratory, Abdominal, Neurological, Skin Soft Tissue, Ent, Orthopaedic, Other Specific, No Specific Source, Prophylaxis, Uncertainty, Not Informative*

*Multiple categories are allowed.*

*When returning your answer, please return a json*

*User prompt:*

*This is the list of indications, return a json with the categories (multiple allowed) for each indication.*

*“abdo pathology”,*

*“sepsis ?hap”,*

*“artholin abscess”,*

*[…]*

The results are then parsed from the JSON-formatted response.

### Appendix S4 GPT4 Token Limits

The number of supplied indications per request is limited by the number of output tokens. We determined that 300 input indications result in ∼3800 return tokens, leaving a 10% safety margin for the 4k return limit.

**Figure S4:**
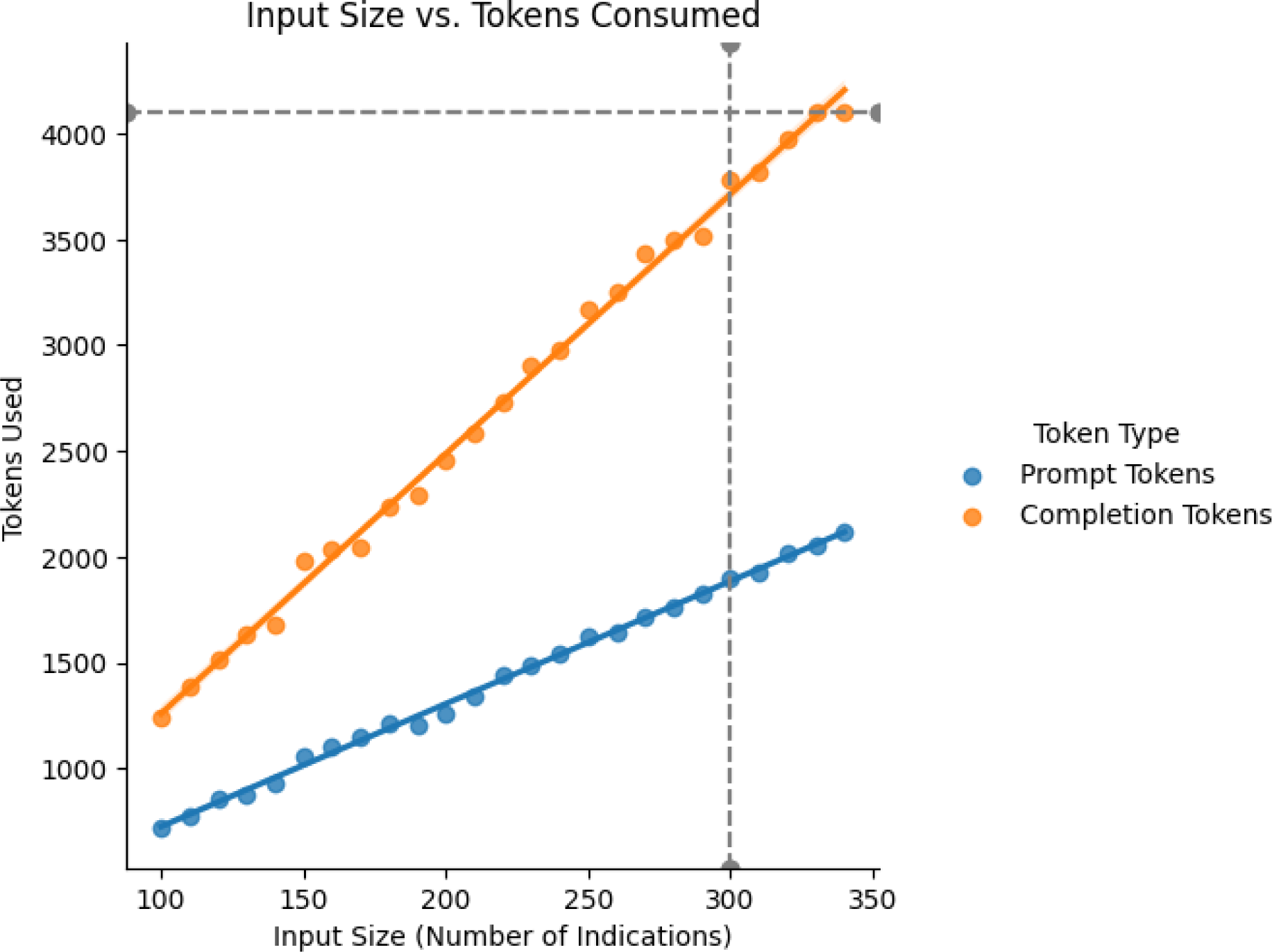
Token consumption (prompt and model output) depending on input size (number of indications.) *Providing a higher number of supplied indications (input to classify) per round is more cost-effective, as the system and user instruction prompts are only submitted once. However, the number of output tokens is limited to 4k or 16k (depending on the model)*.

### Appendix S5 Prescribed Drugs

**Table S5:** Ten most commonly prescribed drugs.

| Drug [Oxford] | Number | Percentage |
| --- | --- | --- |
| Co-amoxiclav | 269945 | 33% |
| Gentamicin | 70002 | 8% |
| Metronidazole | 65094 | 8% |
| Ceftriaxone | 58763 | 7% |
| Flucloxacillin | 47537 | 6% |
| Amoxicillin | 32151 | 4% |
| Ciprofloxacin | 25145 | 3% |
| Vancomycin | 23250 | 3% |
| Piperacillin + Tazobactam (Tazocin equivalent) | 21666 | 3% |
| Clarithromycin | 18607 | 2% |

| Drug [Banbury] | Number | Percentage |
| --- | --- | --- |
| Co-amoxiclav | 39114 | 35% |
| Ceftriaxone | 11369 | 10% |
| Gentamicin | 10251 | 9% |
| Amoxicillin | 7297 | 7% |
| Clarithromycin | 6525 | 6% |
| Flucloxacillin | 5630 | 5% |
| Metronidazole | 4851 | 4% |
| Doxycycline | 4416 | 4% |
| Nitrofurantoin | 3204 | 3% |
| Piperacillin + Tazobactam (Tazocin equivalent) | 2862 | 3% |

### Appendix S6 Prescribing Specialities

**Table S6:** Specialties of the prescribing clinicians, ordered to show the ten most active specialties.

| Clinician Main Specialty [Oxford] | Number | Percentage |
| --- | --- | --- |
| General Surgery | 146719 | 18% |
| Acute General Medicine | 98687 | 12% |
| Trauma and Orthopaedics | 90719 | 11% |
| Acute Geratology | 72371 | 9% |
| Clinical Haematology | 46928 | 6% |
| Obstetrics | 36431 | 4% |
| Neurosurgery | 34666 | 4% |
| Infectious Diseases | 30584 | 4% |
| Plastic Surgery | 29334 | 4% |
| Urology | 29258 | 4% |

| Clinician Main Specialty [Banbury] | Number | Percentage |
| --- | --- | --- |
| Acute General Medicine | 41312 | 37% |
| Acute Geratology | 14912 | 13% |
| Gastroenterology | 9128 | 8% |
| Cardiology | 8682 | 8% |
| Infectious Diseases | 7870 | 7% |
| Trauma and Orthopaedics | 7642 | 7% |
| General Surgery | 6803 | 6% |
| Emergency Medicine | 4924 | 4% |
| Urology | 2885 | 3% |
| Gynaecology | 2287 | 2% |

### Appendix S7 Labelling Coverage

**Figure S7:**
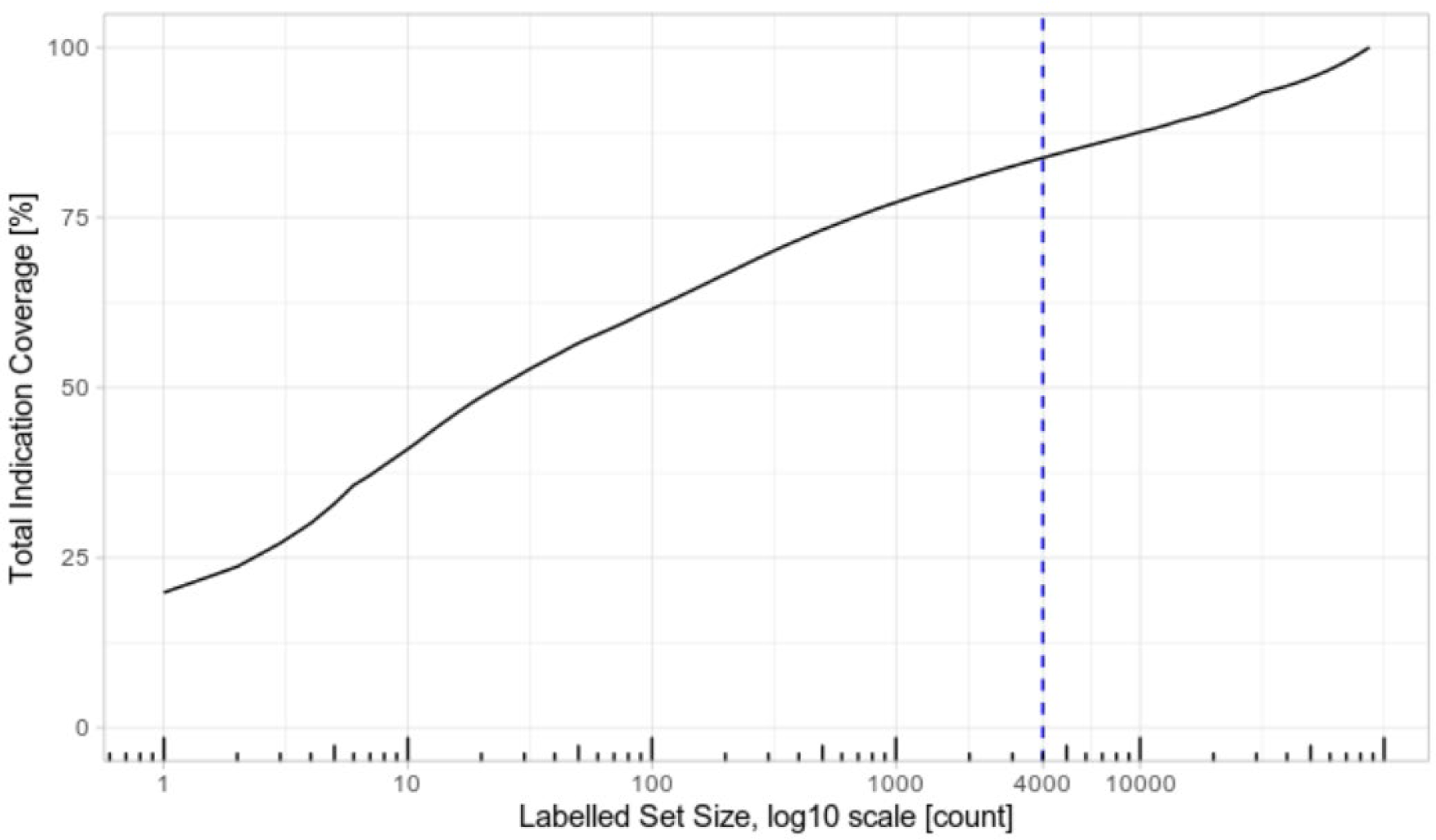
Total coverage of data given a set of labelled data. *Labelling the most common 4000 samples covers 84% of the entire data set*.

### Appendix S8 Uncommon Indications

**Table S8:** Example list of uncommon indications. *Randomly sampled from all indications occurring more than 5 and less than 100 times. Occurrences less than 10 are truncated to <10 for statistical disclosure control*.

| Uncommon Indication | Occurrence | Percentage |
| --- | --- | --- |
| septic left knee | <10 | <1% |
| abdo contamination | <10 | <1% |
| infected femur | 10 | <1% |
| flu prophylaxis | 63 | <1% |
| forearm cellulitis | <10 | <1% |
| mesenteric panniculitis | <10 | <1% |
| intra-abdo inf | 14 | <1% |
| ?dental infection | 14 | <1% |
| cap curb1 | 41 | <1% |
| septic unknown source | <10 | <1% |

### Appendix S9 Class Distributions

**Table S9:**
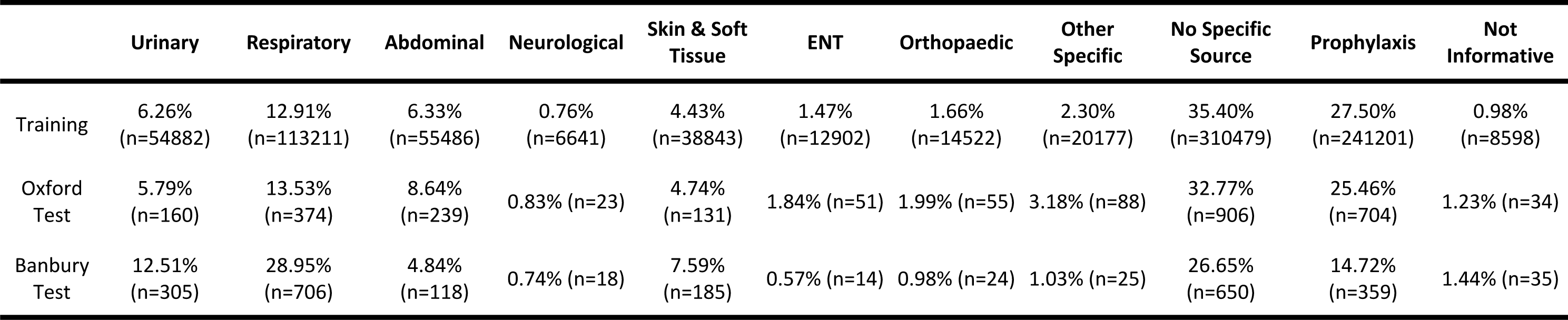
Distributions of the classes within the different data sets.

|  | Urinary | Respiratory | Abdominal | Neurological | Skin & Soft Tissue | ENT | Orthopaedic | Other Specific | No Specific Source | Prophylaxis | Not Informative |
| --- | --- | --- | --- | --- | --- | --- | --- | --- | --- | --- | --- |
| Training | 6.26%<br>(n=54882) | 12.91%<br>(n=113211) | 6.33%<br>(n=55486) | 0.76%<br>(n=6641) | 4.43%<br>(n=38843) | 1.47%<br>(n=12902) | 1.66%<br>(n=14522) | 2.30%<br>(n=20177) | 35.40%<br>(n=310479) | 27.50%<br>(n=241201) | 0.98%<br>(n=8598) |
| Oxford Test | 5.79%<br>(n=160) | 13.53%<br>(n=374) | 8.64%<br>(n=239) | 0.83% (n=23) | 4.74%<br>(n=131) | 1.84% (n=51) | 1.99% (n=55) | 3.18% (n=88) | 32.77%<br>(n=906) | 25.46%<br>(n=704) | 1.23% (n=34) |
| Banbury Test | 12.51%<br>(n=305) | 28.95%<br>(n=706) | 4.84%<br>(n=118) | 0.74% (n=18) | 7.59%<br>(n=185) | 0.57% (n=14) | 0.98% (n=24) | 1.03% (n=25) | 26.65%<br>(n=650) | 14.72%<br>(n=359) | 1.44% (n=35) |

### Appendix S 4 Per Class Prediction Scores

**Table S10.1:** Per class prediction scores on the internal Oxford test set. Values are reported in F1 score, Precision Recall AUC and ROC AUC where applicable.

| Model | Metric | Urinary | Respiratory | Abdominal | Neurological | Skin & Soft<br>Tissue | ENT | Orthopaedic | Other<br>Specific | No Specific<br>Source | Prophylaxis | Uncertainty | Not<br>Informative |
| --- | --- | --- | --- | --- | --- | --- | --- | --- | --- | --- | --- | --- | --- |
| Regex | F1 | 0.87 | 0.63 | 0.63 | 0.68 | 0.76 | 0.54 | 0.3 | 0.11 | 0.78 | 0.93 | 0.20 | 0.00 |
| XGBoost | F1 | 0.64 | 0.81 | 0.9 | 0.76 | 0.83 | 0.83 | 0.7 | 0.71 | 0.91 | 0.96 | 0.66 | 0.9 |
| Base_BERT | F1 | 0.97 | 0.98 | 0.94 | 0.23 | 0.94 | 0.27 | 0.59 | 0.63 | 0.98 | 0.98 | 0.95 | 0.67 |
| Bio_ClinicalBERT | F1 | 0.98 | 0.98 | 0.96 | 0.88 | 0.93 | 0.91 | 0.93 | 0.84 | 0.98 | 0.98 | 0.96 | 0.85 |
| Fine-Tuned GPT3.5 | F1 | 0.98 | 0.97 | 0.95 | 0.93 | 0.83 | 0.8 | 0.95 | 0.77 | 0.97 | 0.97 | 0.92 | 0.99 |
| Few-Shot GPT4 | F1 | 0.98 | 0.96 | 0.83 | 0.88 | 0.87 | 0.79 | 0.87 | 0.30 | 0.34 | 0.94 | 0.78 | 0.30 |
| Regex | PR | - | - | - | - | - | - | - | - | - | - | - | - |
| XGBoost | PR | 0.62 | 0.87 | 0.91 | 0.68 | 0.89 | 0.82 | 0.65 | 0.72 | 0.97 | 0.99 | 0.69 | 0.83 |
| Base_BERT | PR | 0.98 | 0.99 | 0.97 | 0.69 | 0.95 | 0.85 | 0.94 | 0.77 | 0.99 | 0.99 | 0.97 | 0.92 |
| Bio_ClinicalBERT | PR | 0.99 | 1.00 | 0.97 | 0.91 | 0.98 | 0.93 | 0.96 | 0.88 | 0.99 | 1.00 | 0.99 | 0.94 |
| Fine-Tuned GPT3.5 | PR | - | - | - | - | - | - | - | - | - | - | - | - |
| Few-Shot GPT4 | PR | - | - | - | - | - | - | - | - | - | - | - | - |
| Regex | ROC | - | - | - | - | - | - | - | - | - | - | - | - |
| XGBoost | ROC | 0.9 | 0.96 | 0.97 | 0.92 | 0.98 | 0.96 | 0.91 | 0.92 | 0.97 | 0.99 | 0.87 | 0.94 |
| Base_BERT | ROC | 0.98 | 1.00 | 0.98 | 0.97 | 0.99 | 0.96 | 0.99 | 0.91 | 0.99 | 0.99 | 1.00 | 0.98 |
| Bio_ClinicalBERT | ROC | 0.99 | 1.00 | 0.98 | 1.00 | 0.99 | 0.98 | 0.99 | 0.96 | 0.99 | 1.00 | 1.00 | 0.99 |
| Fine-Tuned GPT3.5 | ROC | - | - | - | - | - | - | - | - | - | - | - | - |
| Few-Shot GPT4 | ROC | - | - | - | - | - | - | - | - | - | - | - | - |
| Regex | Accuracy | 0.98 | 0.79 | 0.89 | 0.99 | 0.97 | 0.96 | 0.89 | 0.36 | 0.76 | 0.96 | 0.32 | 0.97 |
| XGBoost | Accuracy | 0.96 | 0.94 | 0.98 | 1.00 | 0.98 | 0.99 | 0.99 | 0.98 | 0.92 | 0.97 | 0.95 | 1.00 |
| Base_BERT | Accuracy | 0.99 | 0.99 | 0.99 | 0.99 | 0.99 | 0.98 | 0.98 | 0.97 | 0.98 | 0.98 | 0.99 | 0.99 |
| Bio_ClinicalBERT | Accuracy | 1.00 | 0.99 | 0.99 | 1.00 | 0.99 | 1.00 | 1.00 | 0.99 | 0.98 | 0.98 | 0.99 | 1.00 |
| Fine-Tuned GPT3.5 | Accuracy | 1.00 | 0.99 | 0.99 | 1.00 | 0.97 | 0.99 | 1.00 | 0.98 | 0.97 | 0.98 | 0.99 | 1.00 |
| Few-Shot GPT4 | Accuracy | 1.00 | 0.98 | 0.96 | 1.00 | 0.98 | 0.99 | 0.99 | 0.94 | 0.64 | 0.96 | 0.97 | 0.96 |

**Table S10.2:**
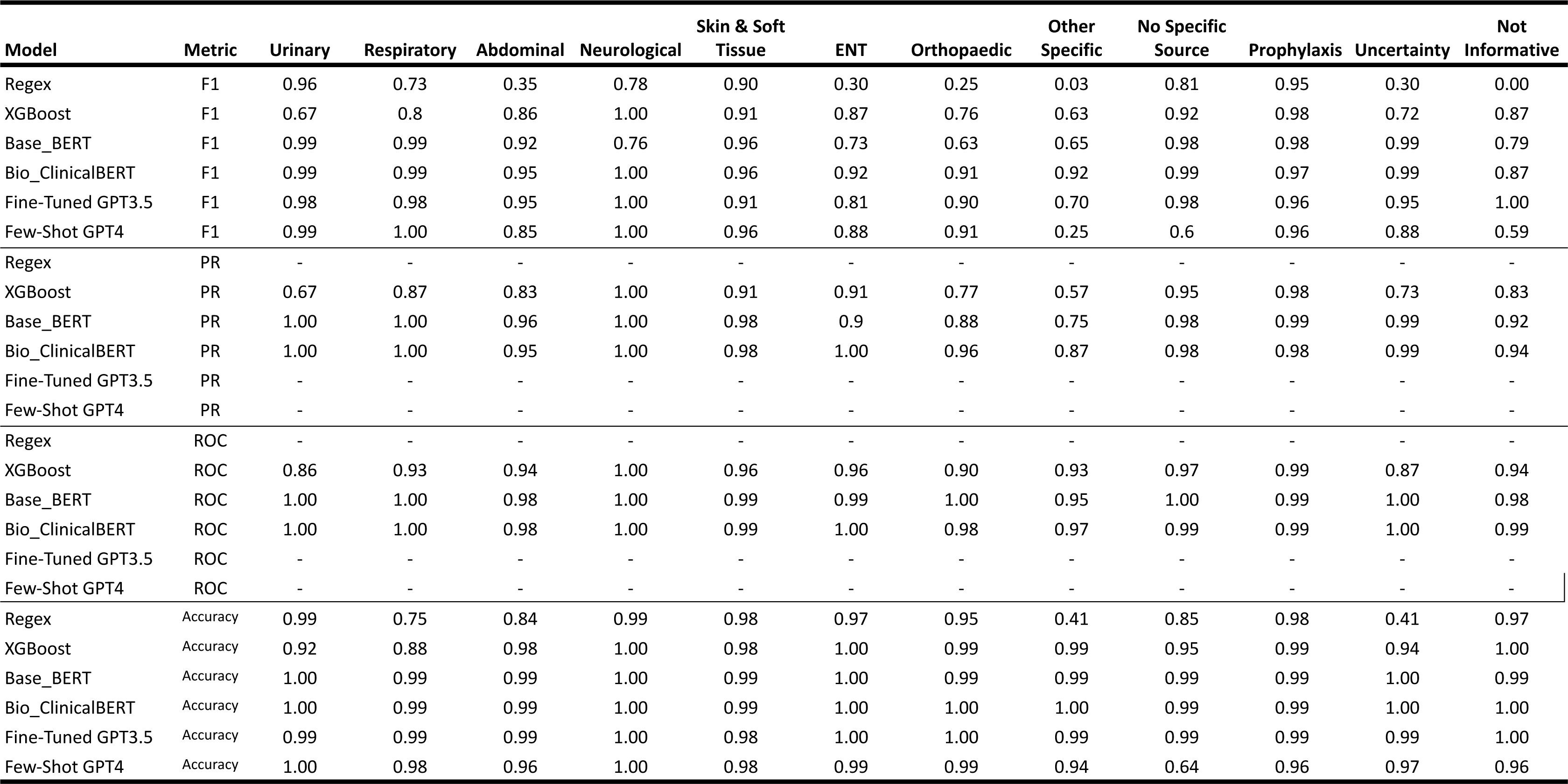
Per class prediction scores on the external Banbury test set. Values are reported in F1 score, Precision Recall AUC and ROC AUC where applicable.

### Appendix S11 Training Set Size Effects

**Figure S11:**
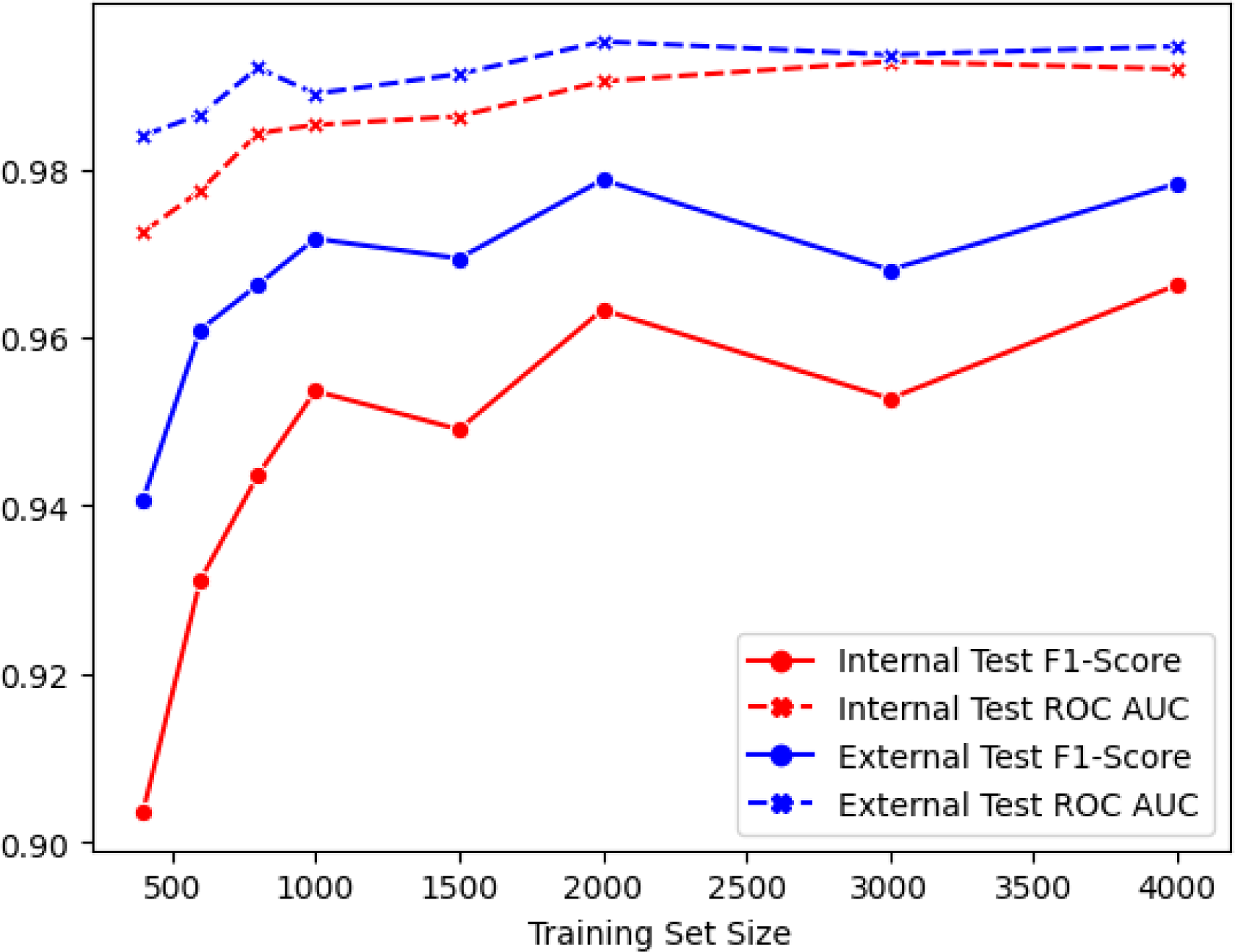
Training set size effect on the performance of Bio+Clinical BERT, evaluated on both F1-Score and ROC AUC. *The training was run on randomly sampled subsets of the training dataset of size [500, 1000, 1500, 2000, 3000, 4000] and evaluated on the same internal and external test sets (2000 samples each)*.

## Notes

### Competing Interest Statement

The authors have declared no competing interest.

### Author Declarations

The National Research Ethics Service South Central Oxford C Research Ethics Committee (19/SC/0403) and the national Confidentiality Advisory Group (19/CAG/0144) gave ethical approval for this work.

